# Skill-level based examination of forearm muscle activation associated with efficient wrist and finger movements during typing

**DOI:** 10.1101/2021.06.09.21258367

**Authors:** Takanori Ito, Yuka Matsumoto, Hayase Funakoshi, Mio Yoshida, Naohiko Kanemura, Takanori Kokubun

**Affiliations:** Department of Health and Social Services, Health and Social Services, Graduate School of Saitama Prefectural University

**Keywords:** Typing, Upper extremity musculoskeletal disorders (UE- MSDs), Motion capture system, Electromyography, Wrist/finger joint movements, Muscle activation

## Abstract

This study aims to elucidate the relationship between the wrist and finger movements and forearm muscle activation of people with different typing skills. It hypothesizes that skilled typists (STs) can move their wrist and finger joints faster than unskilled typists (UTs) can because they can efficiently use their muscles according to the activity characteristics of the flexors and extensors of the wrist joint. During the typing task, we measured wrist and finger movements using a 3D motion capture system and forearm muscle activation using surface electromyography. The angular velocity of the wrist and finger flexion/extension and the muscle activation of the wrist flexors was higher in the STs than that in the UTs while the muscle activation of the wrist extensors was higher in the latter than that in the former. Our results showed that STs may have used their forearm muscles to take advantage of the physical characteristics of the keys and the spring characteristics of their muscles and tendons. It was suggested that they placed less mechanical stress on their finger muscles and tendons when pressing and releasing the keys.

## 1. Introduction

Computers are essential tools in modern society that we frequently use for long periods when working or studying. Most people, regardless of age, gender, or occupation, are required to use computers, with the continuing expansion of digital transformation. Typing is a particularly complex task that relies on many joints and muscles. During typing, finger movements are usually rapid and repetitive and people hold their hands continuously above the keyboard, exposing the tendons and sheaths of their wrist and fingers to mechanical stress (Keller et al., 1998).

Bergqvist et al. (1992) first reported a prospective study wherein typing work induced the risk of upper extremity musculoskeletal disorders (UE-MSDs) of the wrist and finger joints (Bergqvist et al., 1992). In the next largest prospective study, 21% of workers who used computers for more than 15 h/ week developed wrist or finger-related disorders in 1 year, most of which were tendon disorders (Gerr et al., 2002). In a recent review, a dose–response relationship was plotted between the exposure of people to mechanical factors and the risk of UE- MSDs associated with their wrists and finger tendons (Keir et al., 2021). Therefore, it is necessary to investigate how the overuse of wrists and fingers may contribute to the development of UE-MSDs.

In this context, typing is required in bodily kinesthetic learning to avoid overloads on wrist and finger joints and UE- MSDs, such as sports injuries. Previous research on typing proficiency and physical movements has mainly focused on skilled typists (STs) of high skill. Additionally, few studies have assessed the kinematic properties of the wrist and finger joints, even though typing primarily comprises wrist and finger movements (Alizadehkhaiyat and Frostick, 2015, Eygendaal et al., 2007).

To quantify the complex kinematic properties of wrists and fingers during typing, it is necessary to understand their structural characteristics and properly evaluate the coordination between them. The relationship between the synergistic motion of the wrist and finger joints in the sagittal plane is called the tenodesis function (Cabri et al., 2009, Marta et al., 2012). It refers to when passive finger joint flexion is induced during wrist joint extension; and conversely, finger joint extension is induced during wrist joint flexion (Cooney et al., 1989, Horii et al., 1992). Particularly, the muscle tension of the finger joint does not increase excessively and the amount of tendon extension can be increased during wrist joint flexion. In this context, evaluating only the movements of the wrist or finger joint and related muscle activities may lead to a misinterpretation of the mechanical activation of the body. Therefore, the relationship between hand kinematic coordination and muscle activities needs to be properly assessed and the biomechanical risk factors that arise need to be determined. Previous studies have discussed hand and finger kinematics by analyzing the simple tapping of an index finger.

In this study, we evaluated repeated keystroke motions during typing to analyze wrist and finger kinematics during more complex repetitive motions. Movement styles may vary with proficiency because typing is a complex task controlled by many joints and muscles. This study aims to elucidate the relationship between wrist and finger movements and forearm muscle activation for different typing skills. We hypothesized that STs could move their wrists and fingers more efficiently than unskilled typists (UTs) could. The movement of their muscles depends on the characteristics of the flexor and extensor activities at the wrist joint.

## 2. Methods

### 2.1 Subjects

Twelve healthy subjects (nine men and three women) participated in this experiment; they were all right-handed. All the subjects provided written informed consent, according to the Declaration of Helsinki, after receiving a detailed explanation of the purpose of the study and the risks involved. This study was approved by the Ethics Review Committee (approval number: 20,508).

### 2.2 Measurement settings

A PC monitor and QWERTY keyboard (Microsoft, Wired Keyboard 600 ANB-00040, Washington, USA) were placed at a height of 70 cm on a desk. Before beginning the tests, each subject adjusted the chair according to their preferences. We used a 3D motion capture system (Vicon Motion Systems Ltd., Vicon, Oxford Metrics PLC, London, UK; sampling frequency: 100 Hz) and wireless multipoint surface electromyography (EMG) (DELSYS, Delsys Trigno Wireless System, Massachusetts, USA; sampling frequency: 1,000 Hz).

### 2.3 Procedure

We measured the height and weight of each subject and collected the other necessary physical information for the experiment. Infrared-reflecting markers (diameters 4 and 14 mm) were attached to points on the subjects (Fig. 1)— according to the Vicon plug-in-gait full-body (PIG) and wrist and finger joint models (Baker et al., 2007, Cook et al., 2007) —to calculate the sagittal plane of the wrist and the angular velocity of the metacarpophalangeal joint (MCP) of the index finger.

**Fig. 1.**
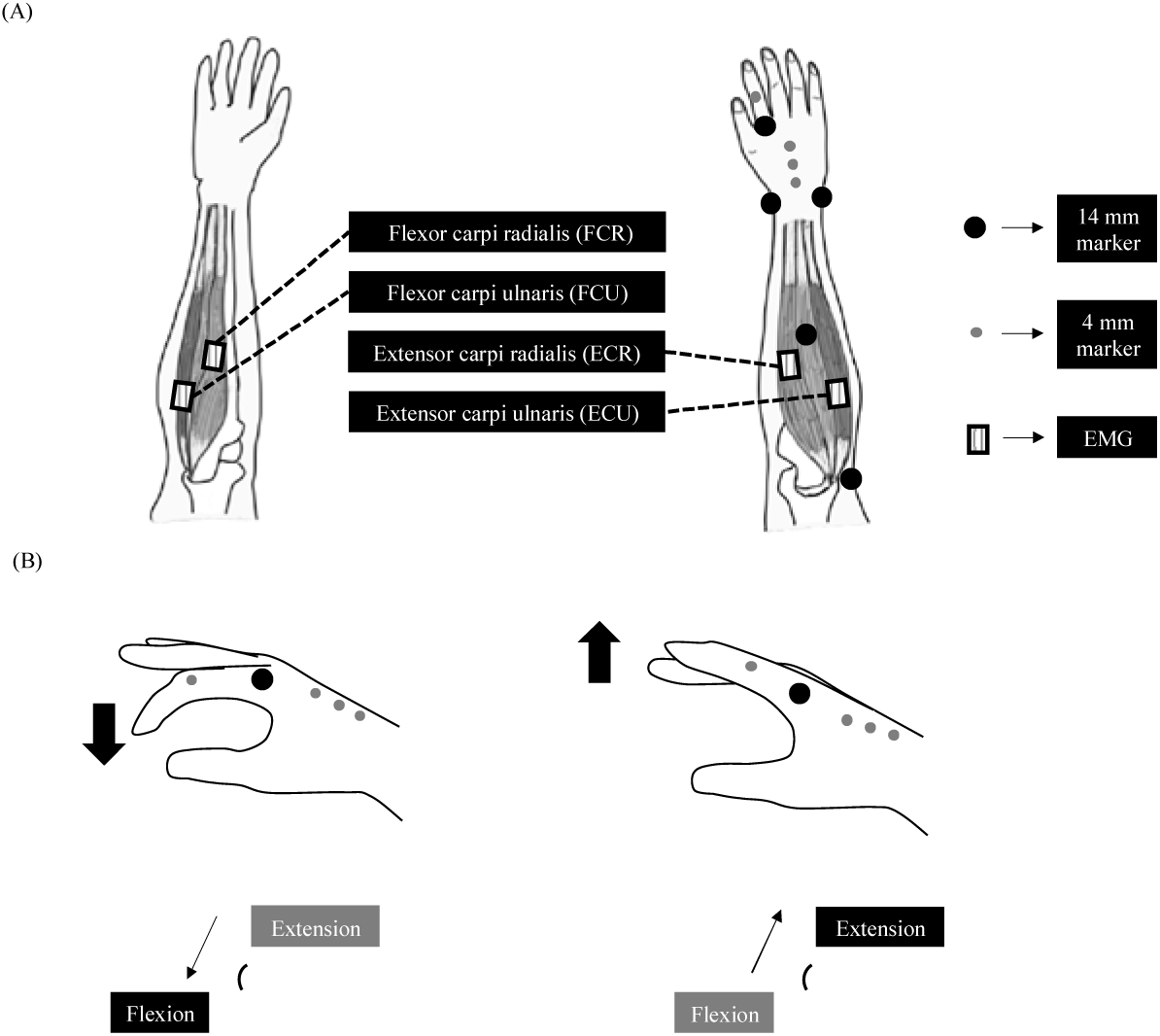
Experimental equipment. (A) Positions of the markers and EMGs (B) Calculation of MCP joint angle

Surface EMG sensors were attached to the right side of the flexor carpi radialis (FCR), flexor carpi ulnaris (FCU), both as the term extensor carpi radialis (ECR), and extensor carpi ulnaris (ECU). The strength of each muscle was measured using maximum voluntary isometric contraction (MVC).

Each subject practiced typing for 5 min before the experiment to become accustomed to the test environment. During the practice trial, each subject adjusted the height of the chair and the angle of the monitor according to their preference. The typing skills of the subjects were pretested on an electronic keyboarding program (e-typing, e-typing Co., Ltd., Aichi, Japan; http://www.e-typing.ne.jp), which had a text display with text that moved as each subject typed.

In the typing test, we instructed the subjects to begin the task after confirming that their hands were placed on the keyboard. All the subjects typed the same sentences—each comprising approximately 5,000 Japanese characters—that were displayed on the monitor. No instructions were provided regarding the typing speed, rhythm, or time limit; all the subjects typed according to their regular style and pace. Each subject performed the task thrice, with short pauses in between.

### 2.4 Data processing

The wrist joint angle was calculated from markers attached to the metacarpal head of the index finger, radial styloid process, ulnar styloid process, and forearm in the model (Fellinger et al., 2010). We expressed that the positive value was the extension and that the negative value was the flexion angle of the wrist joint. The MCP joints were used to define the hand as a rigid body, and the angle of the proximal phalanx segment of the hand on the sagittal plane was calculated and used as the flexion/extension angle (Fig. 1). We expressed that the positive value was the flexion angle and that the negative value was the extension angle for the MCP angle. The 4 mm marker was attached to the enter key on the keyboard to record the typing time and calculate the analysis interval.

The kinematic data were filtered using a fourth-order Butterworth low-pass filter with a cut-off frequency of 6 Hz. After filtering the data, the mean joint angle, changes in the joint angle based on the root mean square (RMS), and joint velocity of the wrist and MCP joint were calculated for each trial.

The EMG data were passed through a fourth-order Butterworth bandpass filter with a cutoff frequency of 20–480 Hz; the filter was full-wave rectified and smoothed at 10 Hz. The maximum voluntary contraction (MVC) normalization technique was applied to the EMG data obtained during the typing sessions.

The 3D motion capture system was used to capture the timing of the first end of the descent of the marker, and the timings of the key input signal and start of the EMG data recording were synchronized in time. The analysis interval was sampled between the first and last entries of the task sentences.

### 2.5 Data analyses

#### 2.5.1 Grouping by the pretest results

All data were obtained using Nexus 2.9 (Vicon Motion Systems Ltd., Vicon, Oxford Metrics PLC, London, UK) and Python 3.7 (Python Software Foundation, Beaverton, Orlando, USA). The accuracy (%) and typing speed (words per minute, WPM) in Japanese for all the subjects in the pretest were obtained, where accuracy is the percentage of number of keys that can be inputted without any mistakes. In a study investigating the keystroke characteristics of both typing novices and experts (Takaoka et al., 2014), both were classified as having a correct response rate of 95% or higher or rates close to it, reaching the required condition for employment.

#### 2.5.2 Kinematic data of wrist and metacarpophalangeal joints

The mean and SD values of the flexion/extension angles of the wrist and MCP flexion/extension angles of the index finger were calculated to analyze the average posture of the wrist and MCP joints during the analysis interval. Similarly, the mean and standard deviation (SD) of the RMS of the data were calculated. The angular velocities of the wrist and index finger MCP joints were investigated by calculating the mean and SD values of the changes in the angles/s .

#### 2.5.3 Analysis of muscle activation in the forearm

To investigate the muscle activation in each forearm muscle, we used amplitude probability distribution function (APDF) analysis for the time series data of %MVC. APDF analysis is a method that expresses the percentage of the total time that the output is below a certain level as the probability of occurrence (*P*) for that output when the EMG is being measured (Jonsson, 1982). Based on these probability values, the muscle activities can be classified into the static activity level, which is equivalent to the activity involved in static posture holding or pausing movement (*P* = 0.1); the average activity level, which is exhibited throughout the movement (*P* = 0.5); and the maximum activity level, which is equivalent to the maximum activity exhibited during the movement (*P* = 0.9). After plotting the function of the power output and appearance probability of each muscle of each subject in each trial, we calculated the mean and SD values of the muscle activation at *P* = 0.1, 0.5, and 0.9.

#### 2.5.4 Identification of press and release times

From analyzing the key input signals of the subjects, we obtained the time at which the input of each key started and ended. The time at which the “U” key was pressed, which is the start of the input, was defined as the press time; and the time at which the fingertip left the “U” key, indicating the end of the inputs, was defined as the release time. The mean and SD values of each data point were calculated and compared between the two groups.

### 2.6 Statistics

After confirming the normality of all the kinematic and muscle activity data using the Shapiro–Wilk test, we investigated the homoscedasticity of the data using the F-test. Since there was no difference in the EMG and kinematic data in each trial (confirmed by one-way ANOVA; *P* < 0.05), the average values of the three trials were used as representative values. The data of the two groups were compared using the unpaired t-test (*P* < 0.05) or Wilcoxon signed-rank test.

## 3. Results

### 3.1 Subjects and environments

The mean accuracy (SD) values in the typing test are shown in Table 1. The mean value of the WPM typed by all the subjects was used as the benchmark for measuring the skill level; subjects who achieved scores higher and lower than the mean were classified into the ST and UT groups, respectively.

**Table 1.** Subjects and Environments

|  | Skilled Typist (ST) | Unskilled Typist (UT) | *p* |
| --- | --- | --- | --- |
| Number | 5 | 7 | — |
| Correct answers(%) | 94.0 (2.3) | 95.1 (2.6) | 0.513 |
| WPM<sup>a</sup> | 219.5 (21.3) | 170.6 (36.2) | *0.033 |
| Age | 21.6 (0.8) | 20.7 (0.5) | 0.050 |
| Height (cm) | 167.3 (5.0) | 166.1 (4.2) | 0.700 |
| Weight (kg) | 63.2 (8.9) | 56.6 (6.4) | 0.202 |
| Hand width (cm) | 7.0 (0.4) | 6.8 (0.1) | 0.466 |
| Finger length (cm) | 10.2 (0.4) | 9.9 (0.3) | 0.305 |
| Chair height (cm) | 42.4 (3.9) | 45.7 (2.8) | 0.127 |
| Screen angle (°) | 129.0(12.4) | 122.9(17.5) | 0.552 |
| Male-to-female ratio | 3:2 | 6:1 | — |
| Right-left handed ratio | 5:0 | 7:0 | — |
| Desktop - notebook PC ratio | 1:4 | 0:7 | — |
| Frequency; PC use times per week | 5.6 (1.0) | 4.86 (1.3) | 0.340 |
| Frequency; PC use times hours per days | 3.8 (1.5) | 2.93 (1.9) | 0.445 |

\* $p < 0.05$ , <sup>a</sup> WPM = Words per minutes

Table.1 Subjects and Environments
|  | Skilled Typist (ST) | Unskilled Typist (UT) | *p* |
| --- | --- | --- | --- |
| Number | 5 | 7 | — |
| Correct answers(%) | 94.0 (2.3) | 95.1 (2.6) | 0.513 |
| WPM<sup>a</sup> | 219.5 (21.3) | 170.6 (36.2) | *0.033 |
| Age | 21.6 (0.8) | 20.7 (0.5) | 0.050 |
| Height (cm) | 167.3 (5.0) | 166.1 (4.2) | 0.700 |
| Weight (kg) | 63.2 (8.9) | 56.6 (6.4) | 0.202 |
| Hand width (cm) | 7.0 (0.4) | 6.8 (0.1) | 0.466 |
| Finger length (cm) | 10.2 (0.4) | 9.9 (0.3) | 0.305 |
| Chair height (cm) | 42.4 (3.9) | 45.7 (2.8) | 0.127 |
| Screen angle (°) | 129.0(12.4) | 122.9(17.5) | 0.552 |
| Male-to-female ratio | 3:2 | 6:1 | — |
| Right-left handed ratio | 5:0 | 7:0 | — |
| Desktop - notebook PC ratio | 1:4 | 0:7 | — |
| Frequency; PC use<br>times per week | 5.6 (1.0) | 4.86 (1.3) | 0.340 |
| Frequency; PC use<br>times hours per days | 3.8 (1.5) | 2.93 (1.9) | 0.445 |

\* $p < 0.05$ , <sup>a</sup> WPM = Words per minutes

The mean (SD) age, height, weight, hand width, finger length, male-to-female ratio, chair height, and screen angle values are presented in Table 1. The ratio of subjects that used desktop to notebook PCs daily was 1:11. The mean (SD) values for the frequency and duration of computer use were 5.2 (1.2) times per week and 3.3 (1.8) h per day, respectively. There were no significant differences between the two groups in any of the data.

### 3.2 Wrist and metacarpophalangeal joint motion

There was no significant difference in the mean extension angle of the wrist joint at the beginning of the typing motion between the two groups. The mean values (SD) of the wrist joint extension angle, RMS, and angular velocity are shown in Fig. 2. The angular velocity of the flexion/extension was predominantly higher in the ST group (*P* = 0.003) than that in the UT group.

**Fig. 2.**
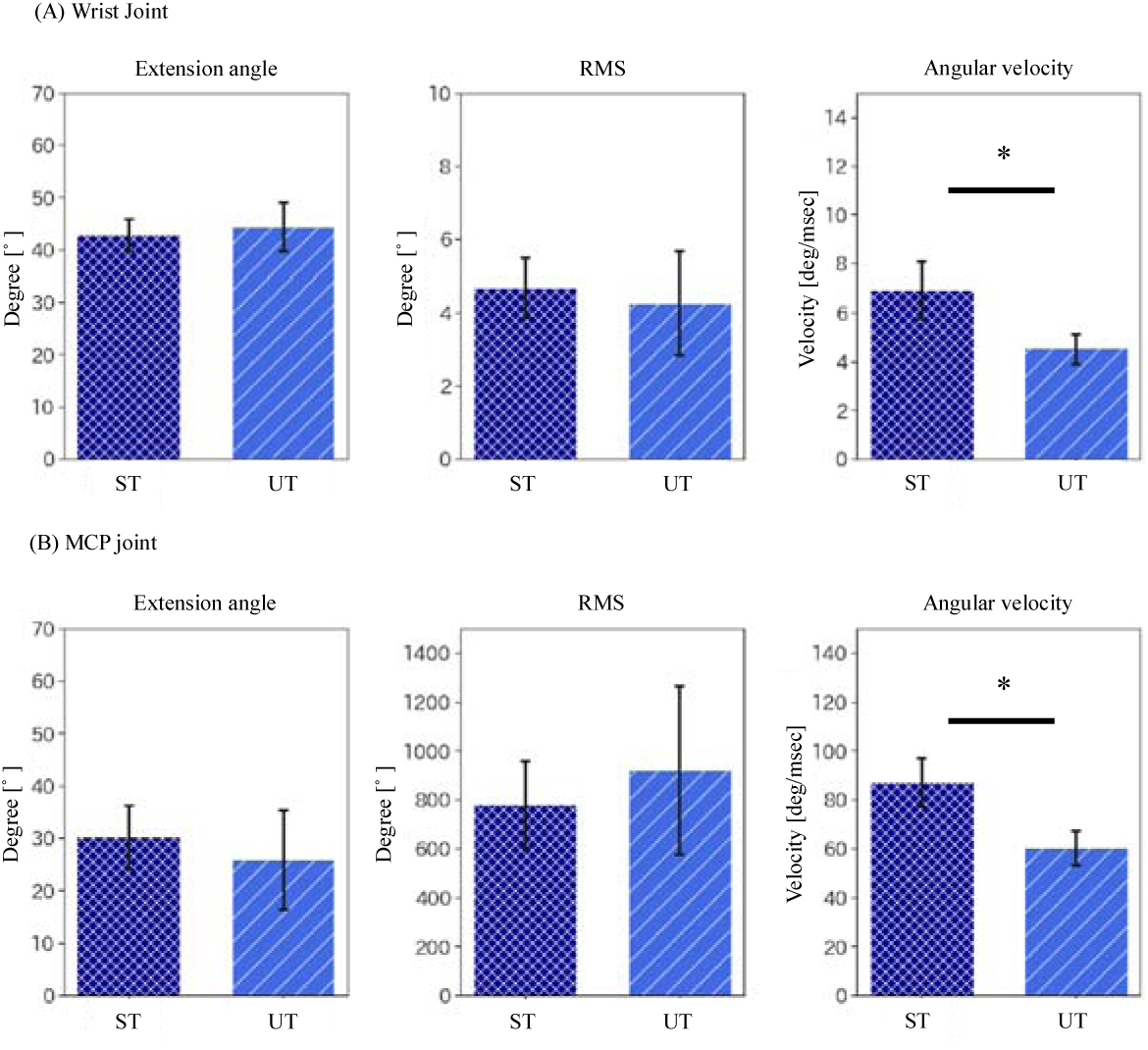
Wrist and metacarpophalangeal (MCP) joint motion. The error bar represents the standard deviation between the subjects. *p* < 0.05, ST = skilled typist, UT = unskilled typist, RMS = root mean square

The mean (SD) values of the flexion angle, RMS, and angular velocity of the MCP joint of the index finger are shown in Fig. 2. The mean angular velocity of the MCP joint in the ST group was significantly higher than that in the UT group (*P* = 0.002). The mean values (SD) of %MVC of FCR, FCU, ECR, and ECU at the static activity level (*P* = 0.1), mean activity level (*P* = 0.5), and maximum activity level (*P* = 0.9) are shown in Fig. 3. The mean %MVC of FCR at *P* = 0.9 was significantly higher than that in the ST group (*P* = 0.023). The mean %MVC of ECR at *P* = 0.1, 0.5, 0.9 were significantly higher than that in the UT group at *P* = 0.040, 0.020, and 0.020, respectively.

**Fig. 3.**
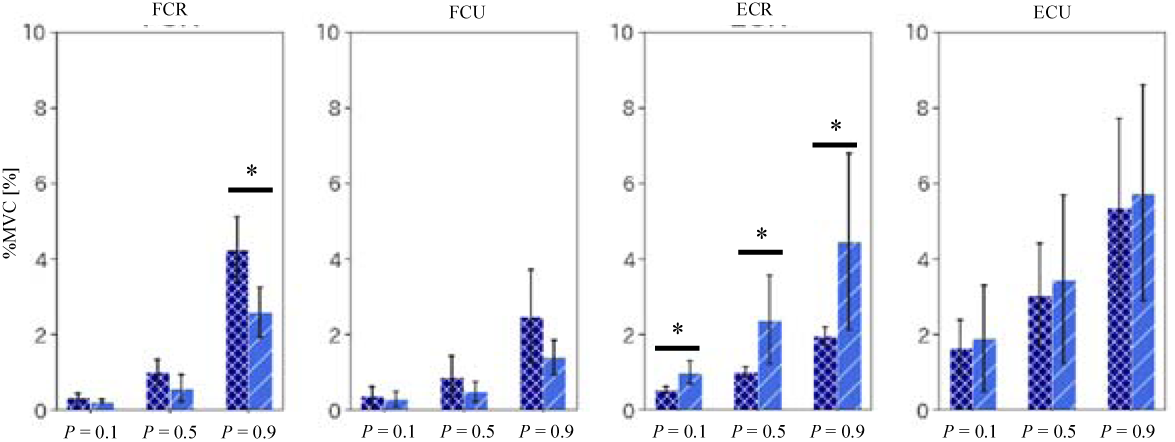
Muscle activation. From left to right, the static activity level (*P* = 0.1), mean activity level (*P* = 0.5), and maximum activity level (*P* = 0.9) are shown. The error bar represents the standard deviation between the subjects. * *p* < 0.05, FCR (flexor carpi radialis); FCU (flexor carpi ulnaris); ECR (extensor carpi radialis); ECU (extensor carpi ulnaris).

### 3.3 Data on press and release time

The means (SD) of the press and release times were 0.12 (0.02) and 0.20 s (0.14), respectively. The press and release times of the ST group were 0.13 (0.01) and 0.11 s (0.01), respectively. The press and release times of the UT group were 0.11 (0.02) and 0.26 (0.15), respectively. There were no significant differences between the two groups. The mean values (SD) of the wrist joint extension angle, angular velocity, flexion angle of the MCP joint of the index finger, and angular velocity during the press and release times are shown in Figs. 4 and 5. The mean angular velocity of the wrist joint extension during the release time in the ST group was significantly higher than that in the UT group (*P* = 0.005). The mean value of the angular velocity of the MCP joint flexion/extension of the index finger during the press and release times in the ST group was significantly higher than that in the UT group (*P* = 0.002, 0.006).

**Fig. 4.**
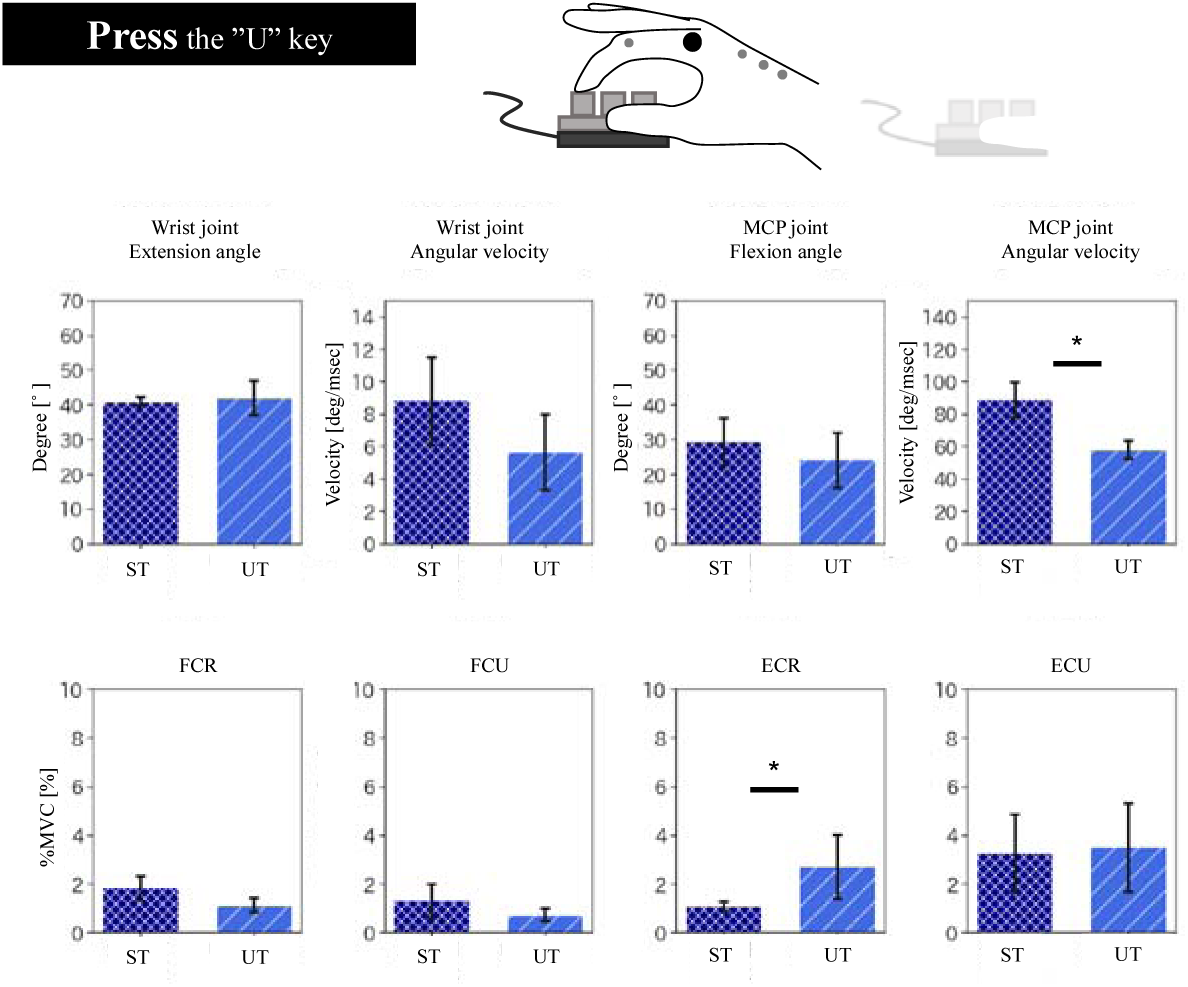
Press time data. Data on when the “U” key is pressed. From left to right, the wrist joint extension angle, angular velocity, flexion angle of the MCP joint of the index finger, angular velocity, and %MVC of the FCR, FCU, ECR, and ECU are listed. The error bar represents the standard deviation between the subjects. * *p* < 0.05

**Fig. 5.**
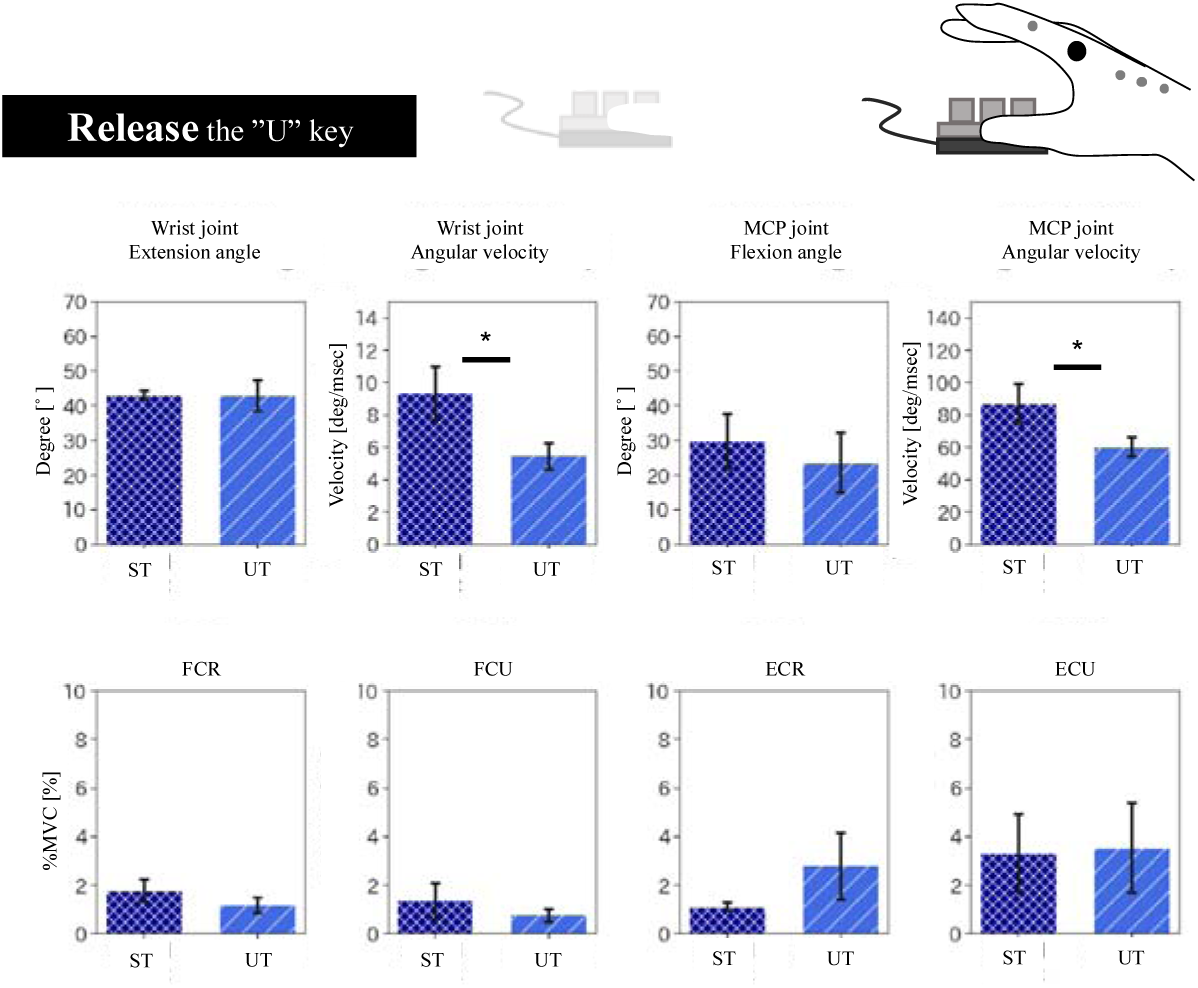
Data on release time. Data on the time between releasing the “U” key and the start of the next keystroke. From left to right, the wrist joint extension angle, angular velocity, flexion angle of the MCP joint of the index finger, angular velocity, and %MVC of FCR, FCU, ECR, and ECU are listed. The error bar represents the standard deviation between subjects. * *p* < 0.05

The mean values (SD) of the %MVC of the FCR, FCU, ECR, and ECU during press and release are shown in Figs. 4 and 5. The mean values (SD) of the %MVC of the ECR during the press time of the ST group were significantly lower than those of the UT group (*P* = 0.020).

## 4. Discussion

We hypothesized that the subjects of the ST group could move their wrist and finger joints faster than those of the UT group could because they could use their muscles according to the activity characteristics of the flexors and extensors of their wrist joint. The ST group had a higher muscle activation at the maximum activity level of the FCU, whereas the UT group had a higher muscle activation at all the ECR activity levels. The subjects of the ST group also moved the MCP joints of their wrists and index fingers faster than those of the UT group did. The press and release time results clearly explained the muscle activation and kinematics of the subjects during the experiment. This study revealed that repetitive keystrokes with muscle activity controls for efficient wrist and finger movements reduced muscle activation during typing. This study emphasizes the significance of assessing not only muscle activity but also the complementary movements of the wrists and fingers simultaneously when considering muscle activation and the risks associated with UE-MSDs during typing.

### 4.1 Muscle activation and movement during experiment

The maximum activity level of the FCR (*P* = 0.9) was higher in the ST group. This value is an index of the peak level of muscle activity involved in the task performance (Szeto et al., 2009); it refers to the muscle activity acting on the pressed keys during typing.

Key presses are coordinated by the muscle activities of the wrist and finger flexors (Dennerlein et al., 2007). Additionally, because the finger flexors were stretched in the wrist extension posture, the mechanical stress on them and the tendons may have increased if active tension was generated in the stretched state when pressing the keys. Therefore, the controlled muscle activity of the ST group may be attributed to the increase in the peak level of muscle activity in the wrist flexors, thereby decreasing the muscle load and mechanical stress on the finger flexors.

All the ECR activity levels (*P* = 0.1, 0.5, and 0.9) were higher in the UT group. Static activity level is an index of muscle activity related to maintaining a static posture (Simoneau et al., 2003); it was inferred that the muscle activity was inefficiently controlled due to static posture and the highly antagonistic action of the finger flexor when pressing the keys.

### 4.2 Kinematics of wrist and finger joints during experiment

No significant differences were observed in the extension angle and RMS of the wrist between the ST and UT groups. However, the angular velocity of flexion/extension was higher in the ST group than that in the UT group. The kinematic parameters of the MCP joint of the index finger showed no significant differences in the joint angle and RMS. The higher typing speed of the ST group was due to the fast movements of the wrist and finger joints and coordinated fast actions of the joints.

To move the wrist and finger joints efficiently, it is important to convert variables, such as inertia and gravity, into joint movements (Dennerlein, Kingma, 2007, Qin et al., 2014). As mentioned in 4.1, the ST group had muscle coordination that increased and decreased the FCR and ECR muscle activities, respectively. This may have allowed the ST group to maintain the downward inertial force generated in the fingertip immediately before pressing the keys and reduced the braking action. As a result, the joint angular velocity of the wrist and finger when pressing the keys was higher in the ST group.

Additionally, the ST group performed muscle activity control to reduce the ECR muscle activity during the experiment. This indicated that the ST group effectively used the spring action of the finger flexors to support the movement of the fingertips in resisting gravity during the transition from pressing the keys to releasing them. This inference also indicated the existence of a relationship between the fast movements of the wrist and finger joints.

### 4.3 Muscle activation and movement when pressing and releasing keys

We evaluated the muscle activation of the wrist joint during press and release times. The types and combinations of fingers that a person uses while typing vary by skill; however, the number of typists that do not use the index finger is small (Feit et al., 2016). Additionally, the “U” key was most frequently entered using the right index finger, because vowels (a, e, i, o, u) in Japanese are always entered after the consonants.

The two groups showed similar activations of the wrist flexors (FCR and FCU) during the task and muscle activities during the press and release times. The ST group was also able to perform finger and wrist joint movements faster than the UT group could. Since the flexion movements of the MCP joint during the typing movements were synergistic with those of the wrist joint (Dennerlein, Kingma, 2007), it can be assumed that the ST group took advantage of the excessive muscle activity of the wrist flexors during keystrokes. Additionally, because the fingertips were in high-speed contact with the keys when pressing them, the wrist flexors may have acted excessively to maintain the posture of the hand against the reaction force from the keys. Finger flexor activity is activated after the onset of downward finger movement (Kuo et al., 2006); the ST group may have activated their wrist flexors in a coordinated manner. The ST group used this strategy because it increased the kinetic energy of their fingertips immediately after pressing the keys, producing a key release, which facilitated keystroke switching.

Wrist posture affects muscle strength and activation patterns (Visser et al., 2000), and sustained low-intensity muscle activity has been associated with wrist extension postures during typing (Dennerlein and Johnson, 2006) with the wrist extensor muscle activity decreasing as the extension angle of the wrist decreases (Simoneau, Marklin, 2003), possibly because of changes in the muscle length and moment arm.

In this study, there was no difference in the angle of the wrist joint or the degree of change in the angle between the two groups; however, the muscle activation of the static activity level of the ECR, an extensor muscle of the wrist joint, was significantly higher in the UT group than that in the ST group. This may be due to the over-activation of the ECR in the UT group due to the lack of minimal muscle activity control despite the longer moment arm of the muscle. This action is antagonistic to the flexion movement of the finger and, therefore, may increase the muscle load on the finger flexors when pressing the keys.

Furthermore, the ECU muscles cause the hand to be raised after pressing the keys (Dennerlein, Kingma, 2007). The UT group could not efficiently utilize the spring properties of the finger flexor tendons, resulting in the overuse of the muscles acting on the extensor muscles of their wrist joint when lifting their fingertips. This may have increased the EMG load on the ECU muscles when releasing the keys. Additionally, there is a proportional relationship between the distance of the spatial movement of the hand after key release and the time interval before the next keystroke (Feit, Weir, 2016). Therefore, the UT group, which tended to have longer key release times, may have increased the muscle load on the ECU muscles during key release because of excessive attempts to lift and move the hands.

### 4.4 Limitations

The measurement conditions in this study were not strict. The postures of the subjects were not constrained during the test; they could place their forearms on the desk or keep them floating. Since placing the forearm on the desk may decrease the activation of the extensor muscles of the wrist joint, the distance between the upper body and the keyboard and the space on the desk in front of the keyboard needs to be checked at the beginning of the measurement; and the effects of differences in the contact time and contact area between the desk and the forearm on the body during the task should be determined.

In this study, we examined the differences in the physical parameters of skill levels among healthy adult students. Therefore, the findings of this study cannot be generalized completely.

The press time and release time were used as the analysis intervals, and the index finger movements were analyzed thoroughly. However, we could not confirm which finger each subject used to input the “U” key. Thus, we may not have been able to exclusively evaluate the movement of the index finger.

Furthermore, because we did not measure the force exerted by the fingertip during keystrokes, we could not demonstrate a direct relationship between the angular velocity of the MCP joint and the actual force generated during keystrokes. In future investigations, we intend to evaluate the movements of fingers other than that of the index finger and examine the relationships between changes in the posture, muscle activation, and the force exerted by fingertips during specific keystrokes.

## 5. Conclusions

The velocity of the wrist and finger joints and muscle activation of the forearm during typing differed according to the skill level of the subjects, and it was inferred that these differences were caused by using muscles during keystrokes.

This study emphasizes the significance of assessing not only muscle activity but also the complementary movements of wrists and fingers simultaneously when considering muscle activation and the risks associated with UE-MSDs during typing.

## Data Availability

The data are not publicly available due to their containing information that could compromise the privacy of research participants

## Conflicts of Interest

The authors declare that there are no conflicts of interest.

## Notes

### Competing Interest Statement

The authors have declared no competing interest.

### Funding Statement

There is no applicable funding.

### Author Declarations

All participants in this study provided written informed consent, following a detailed explanation of the purpose and risks according to the Declaration of Helsinki. This study was approved by the ethics review committee at Saitama Prefectural University, Saitama, Japan (approval number: 20,508).

